# Compartmental Model for Epidemics with Contact Tracing and Isolation under Arbitrary Degree Distribution

**DOI:** 10.1101/2024.05.15.24307402

**Authors:** Elan Ocheretner, Amir Leshem

## Abstract

The recent COVID-19 epidemic demonstrated the need and importance of epidemic models as a tool for policy-making during times of uncertainty, allowing the decision-makers to test different intervention techniques and scenarios. Furthermore, tools such as large-scale contact tracing became technologically feasible for the first time. While large-scale agent-based simulations are nowadays part of the toolboxes, good analytical models allow for much faster testing of scenarios. Unfortunately, good models that consider contact tracing and quarantine, and allow for different degree distributions do not exist. To overcome these shortcomings of existing models we propose a new simple compartmental model that integrates quarantine and contact tracing into the SIR compartmental models with arbitrary degree distribution of nodes to better understand the dynamics of the disease under various parameters of intervention and contagion. Consequently, we analytically derive the epidemic threshold as a function of the degree distribution and the model parameters when both quarantine and contact tracing are used. Simulation results demonstrate and quantify the benefits of quarantine and contact tracing and show the effectiveness of such measures over a large range of epidemic parameters.

## Introduction

The outbreak of COVID-19 affected people around the world and provided health, social, and economic challenges [1]. Estimating the impact of the disease [2, 3] and use of mitigation measures [4, 5] can help shape the effect on the population and combat the outbreak.

Epidemiology is the field of science for mathematical modeling for such problems [6, 7]. When researching the field of epidemiology, we can see two distinct approaches: agent-based simulation [8, 9] and compartment modeling [10–12]. The advantage of agent-based simulation is that you can tailor the epidemic to a specific case by simulating different disease parameters on a certain population on the individual level. However, there is a severe limitation with the agent-based simulations. This approach is computationally expensive and time-consuming, as discussed in [13]. This makes it difficult to conclude anything general regarding the epidemic in a given population without simulating the model for many repetitions. The compartmental approach proposes compartmentalizing the population in different states and devising ordinary differential equations to describe at what rate individuals travel between different compartments [14]. The compartment models offer an easy, computationally inexpensive way to model the propagation of the disease and provide insight for future scenarios, allowing quick decision-making.

The most popular compartmental model is the SIR model, it divides the population into three models S—susceptible, I—infected, and R—recovered. Examples of other models include SIS (susceptible—infected-susceptible) [15] where infected individuals become susceptible again after some time, SIRQ (susceptible—infected—recovered—quarantined) [16] which incorporates quarantine of infectious individuals. Usually, when such models are used, it is assumed the population is a well-mixed one. The issue with such an assumption is that it doesn’t describe reality [17]. To resolve this a degree-based approach is used [18, 19], and it assumes that vertices with the same degree behave similarly, in [12] we see that this approximation is a good one. [20–22] solve the case of the SIR model with degree distribution, where [22] specifically explores the subject on a more complex network.

In the recent COVID-19, we saw that implementing contact tracing and isolation techniques could be used to control the outbreak [23], [24], [25], [26], [27], [28], that is why in our research, we focused on incorporating quarantine and contact tracing into the classic SIR compartmental model under any arbitrary degree distribution. This grows more relevant in time as improvements in technology can help make contact tracing easier [29]. Some papers try to solve this problem in the agent-based approach [30], [31], [27], while others used compartment models but only using fully-mixed models [32], [33]. Our model enriches the SIR model by changing S, and I’s compartments to two compartments each (Not quarantined and quarantined states for each previous state) and add additional transitions between states. Papers [30], [34], [32], [33] are closest to our model. The model in [33] is fully mixed and does not address any social networks. [30] quantifies the network effects of contact tracing using configuration models, but doesn’t provide the equations of the model for further research. In addition, [30] and [34] assume a different tracing process – the tracing occurs immediately as an infected individual enters quarantine. The rate of contact tracing differs in several methods of tracing, as can be seen in [29], also the delay highly affects the impact of contact tracing [35]. In our model, we assume there is a delay to contact tracing similar to [32]. In our research, we simplify and provide easier equations to work with than in [32]. We devise a simple, equation-based way to quantify the effect of contact tracing and isolation on epidemic spread. For the case of a regular graph, we provide an analytical solution to reduce the equations to a single equation to solve to further simplify the calculation. In addition, we show that the regular graph case is similar to Erdős–Rényi. This important result means that any degree distribution that can be approximated as Erdős–Rényi and only one equation needs to be solved. Moreover, we can clearly see in the results the validity of our model and the effectiveness of tracing and quarantine measures over a large range of epidemic parameters. In the future, our model can be a foundation for other models incorporating any additional measures of epidemic control.

## Results

In this paper, we extend the classical SIR models with arbitrary degree distribution [12], [18,19], to the case where quarantine and contact tracing are available measures to limit the spread of the epidemic. These measures have been widely applied during the recent pandemic [36–39], however, we were unable to find simple compartmental models incorporating these measures. Our main results include the development of the compartmental model incorporating both quarantine and contact tracing, for an arbitrary degree distribution model. The model is depicted in Fig. 1. For this model, we derive a simple closed-form formula for the epidemic threshold.

**Fig. 1:**
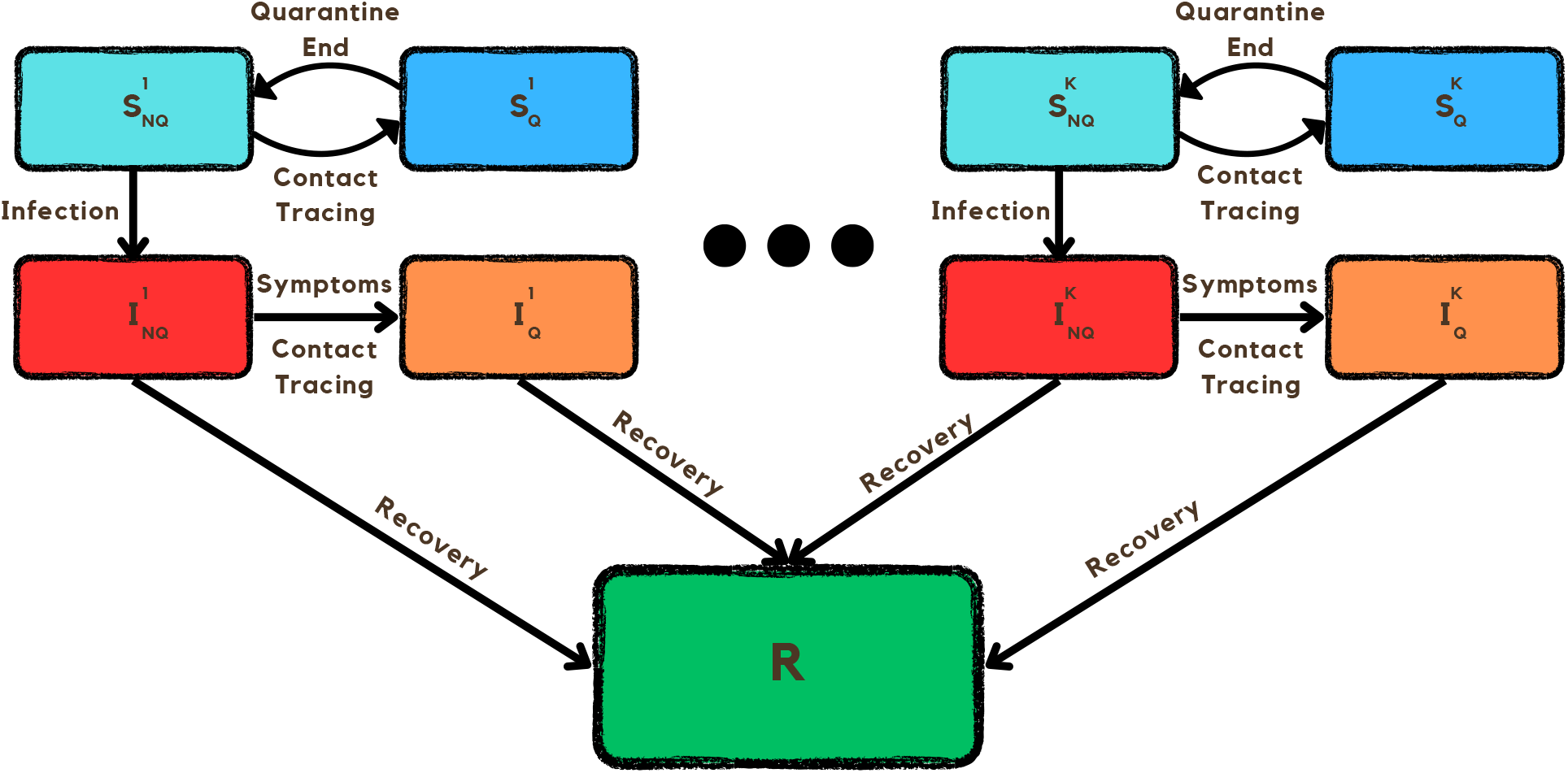
An illustration of the SIRCQ model proposed in this paper. For each degree *k*, we have four possible compartments 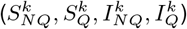, whereas the recovered compartment *R* is for all degrees.

Then we significantly simplify the equations in the case of a contact graph with a fixed degree of all nodes. This generalizes the classical SIR model, where each node can be connected to all other nodes. We show, by simulations, that this simplification provides an excellent approximation for the Erdős–Rényi family of random graphs [40], [41].

To validate our theoretical results, we performed extensive comparisons with agent-based models for both scale-free degree graphs and regular graphs. First, we compared the temporal epidemic evolution, and then we evaluated the proportion of infected people under various model parameters. We also validate the formula for the epidemic threshold in both cases. The results show good agreements between theory and simulation and validate our results.

### SIRCQ model

We begin with a detailed description of the proposed model. Note that the interplay between contact tracing and quarantine is complex and is further complicated by the stochastic nature of contact tracing and the variability in symptom onset. Our model aims to capture these dynamics to provide insights into the effectiveness of quarantine and contact tracing as strategies to control the spread of infectious diseases. The model takes the classical SIR compartmental model with degree distribution and splits each susceptible (*S*^*k*^) and infected (*I*^*k*^) compartment into two compartments each, one with quarantined individuals and another with non-quarantined individuals. In total, we have five distinct compartments for each degree in the contact graph: susceptible individuals who are quarantined 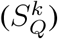, susceptible individuals who are not quarantined 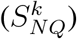, infected individuals who are quarantined 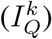, infected individuals not quarantined 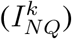, and recovered individuals (*R*). Fig. 1 depicts the transitions between compartments.

While transitions are made between compartments of each degree, the transition rate is determined by the cumulative amount of infected individuals in all infected compartments. The dynamics of the model are governed by a set of differential equations that account for the arbitrary degree distribution of the contact network, representing the heterogeneity in the number of contacts per individual. For each degree k, there are five equations corresponding to the five compartments. These equations represent the net inflow of individuals to the corresponding compartment and are coupled through the total number of infected individuals.

We will start describing the transitions in the compartments with the infection process, which moves individuals from 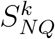 to 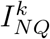. Newman [12] presents the Susceptible — Infectious — Recovered (SIR) model with degree-based distribution. The excess degree distribution is used in order to describe the infection process. We will use a similar process for our model. The reason excess degree distribution is used is that a susceptible vertex (*A*) can be infected by an infected neighbor (*B*) only if *B* has another infected neighbor (*C*) who is not susceptible. Thus, to get the infection probability we need to consider the excess degree distribution of *B*, so that we don’t include *A*. In short, the infection rate of each susceptible individual depends on 3 parameters: the probability of infection upon contact, the degree of the individual, and the probability that their contact/neighbor is infected, which as we mentioned above is determined by the excess degree of the contact.

Symptomatic transfer to quarantine is another feature of our model, where individuals in the 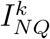 compartment are moved to the corresponding 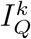 compartment based on the onset of symptoms. This process considers the delay between infection and the onset of symptoms, which plays a significant role in the disease dynamics. In our model, symptoms are assumed to follow a geometrical probability distribution over the course of the infection. This process considers the delay between infection and the onset of symptoms, which plays a significant role in the disease dynamics.

Contact tracing is modeled as a mechanism that transitions individuals from the 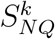 to the corresponding 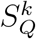 compartment, and from the 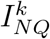 to the corresponding 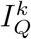 compartment, upon identification of their contact with a *known* infected individual, which means the traced individual has to be from IQ. The efficiency of the contact tracing process is a critical parameter, influencing the rate at which individuals are quarantined and thereby affecting the spread of the infection. We parameterize the efficiency of contact tracing as the rate of contact tracing, which is assumed to follow a geometrical probability distribution from the time an individual enters the 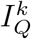 compartment. This can represent different contact-tracing mechanisms, such as electronic contact tracing and epidemiological investigations [29, 35]. Similarly to infection, contact tracing is affected by the excess degree distribution (which is a function of all infected compartments) as the infected and quarantined individuals had to be traced from another neighbor, who is in quarantine, and is not necessarily of the same degree.

The recovery process is modeled through a transition from both IQ and INQ compartments to the R compartment, using a geometrical probability distribution similar to the symptoms’ onset. The recovery signifies the development of immunity or the end of the infectious period.

The combination of all this information is depicted in Fig. 1, we can see all the compartments for each degree and all the possible transitions mentioned above. This model provides a quick and valuable tool for public health officials and policymakers to evaluate the implications of different intervention strategies, offering a more nuanced understanding of epidemic control in the face of uncertainty and incomplete information. The implications of our findings are particularly relevant for the strategic design of measures to mitigate the spread of future epidemics. Detailed derivation of these equations is given in the methods section.

### Epidemic threshold of SIRCQ model

Our main theoretical result is the computation of the epidemic threshold for the SIRCQ model in two cases: Arbitrary degree distribution and for the special case of a regular graph. The computation utilizes the technique of [42] by defining the next-generation transmission and transition matrices to calculate *R*_0_. For arbitrary degree distribution, the epidemic threshold is growing linearly with the infection parameter and the mean degree of a neighbor, also known as the mean excess degree and inversely proportional to the sum of the inverse of the mean time for the appearance of symptoms and the recovery rate. The detailed calculation is given in the Methods section under “Calculation of the epidemic threshold for arbitrary degree distribution”.

The mean degree of a neighbor is the important parameter, similar to [12] for the degree-based SIR. As is expected, contact tracing does not affect the epidemic threshold, but rather reduces the spreading rate when the epidemic evolves. The logic behind it is that when everyone is virtually susceptible and there are no infected people in quarantine, contact tracing is not yet effective and does not determine the disease outbreak. We also provide a simpler derivation for the case of a k-regular graph. This generalizes the standard SIR case by limiting the number of contacts of each person to a specific number.

### Comparison of the compartment model and agent-based model

To validate our model we compare the numerical solution of the compartmental model equations with an agent-based model (ABM). The results show good agreement between the models, which verifies our equations. Fig. 2 compares the temporal behavior of the two models for power law distributed degrees of the nodes. The minimal degree was 8 and the mean degree was 40. This generates a highly heterogeneous degree distribution. The disease parameters are determined by the mean transition time between compartments that are geometrically distributed. The probability of infection per contact (*β*) was selected to be 0.02, the recovery rate (*γ*) was chosen to be 0.14 (7 days), the contact tracing rate (*p*_*ct*_) was selected to be 0.5 (2 days), the rate of return of susceptible individuals from quarantine (*θ*_*s*_) was chosen to be 0.07 (14 days), and the rate of symptoms appearance (*p*_*symptoms*_) was selected to be 0.25 (4 days).

**Fig. 2:**
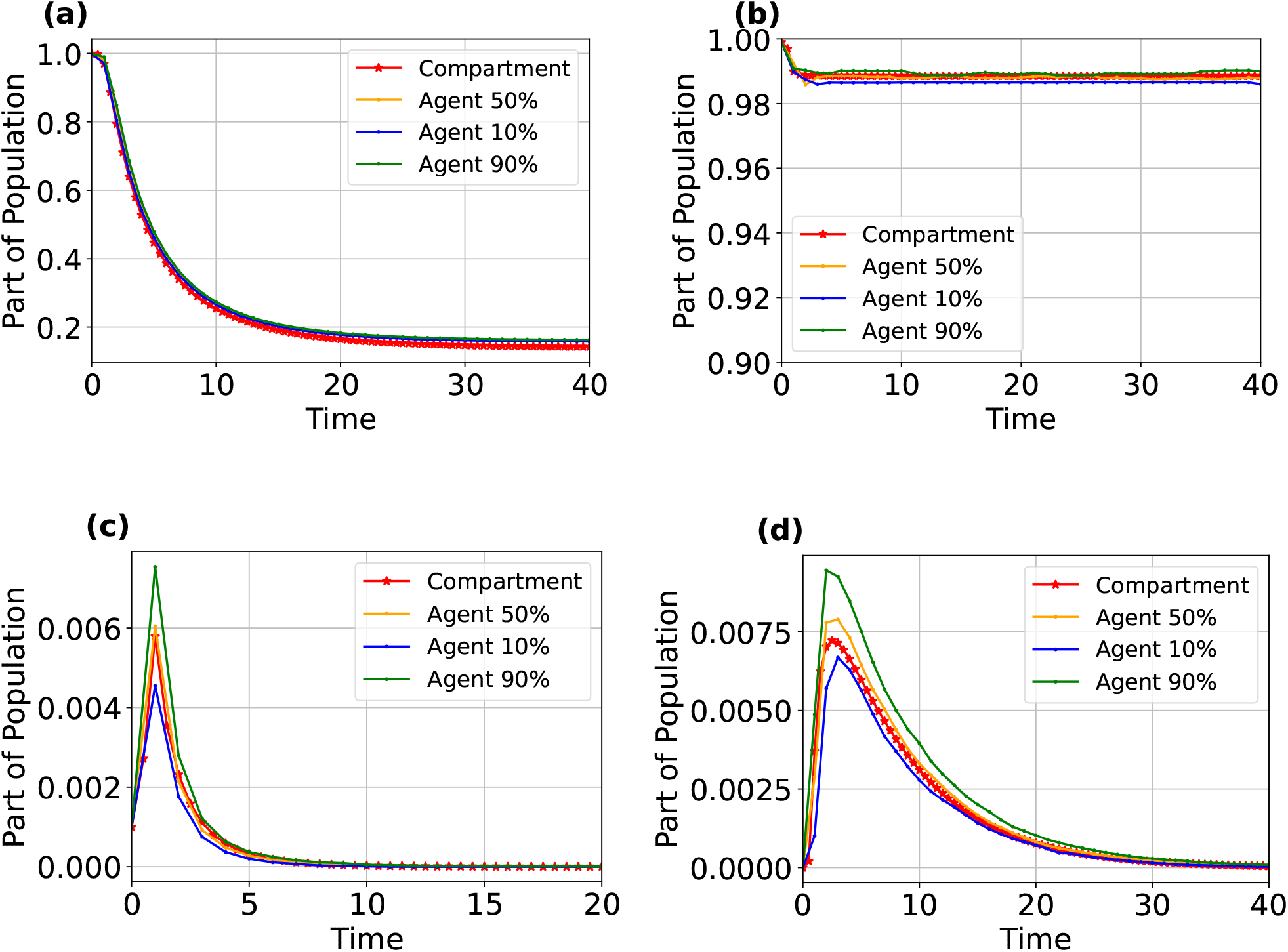
Comparison of the compartment model and agent-based model in the power law case. The sub-figures are susceptible in Newman’s model (a) susceptible in the SIRCQ model (b), infected and non-quarantined (c), and infected and quarantined (d). The range is between 0 and 1 because we normalize by the population. The parameters of the epidemic are: *β* = 0.02, *γ* = 0.14, *p*_*ct*_ = 0.5, *θ*_*s*_ = 0.07, *p*_*symptoms*_ = 0.25. For (d) symptoms and contact tracing are zero (*p*_*ct*_ = 0, *p*_*symptoms*_ = 0).

Fig. 2a shows the relative population of the susceptible compartment as a function of time in Newman’s model (no quarantine and no contact tracing), The reproduction number in this case is *R*_0_ ≈ 40. If we add the symptoms’ onset and contact tracing we get Fig. 2b. We can clearly see different behavior in the susceptible population, the proportion of infected people in the population after the epidemic ends (*r*_∞_) is lowered from 0.86 to 0.012. This directly expresses the power of quarantine and contact tracing, showing how important these measures are.

Fig. 2c and Fig. 2d show the relative infected population as a function of time. In all the results in Fig. 2 we can see a clear agreement between the results of the agent-based and compartmental models. The compartmental model is close to the median of the agent-based model and falls well within the 10% − 90% confidence interval.

Similarly, Fig. 3 presents the same results for regular graphs. In this case, there is also a clear agreement between the results of the agent-based and compartmental models. This further enhances the validity of our results by showing that the compartmental model agrees with the agent-based model for two significantly different degree distributions.

**Fig. 3:**
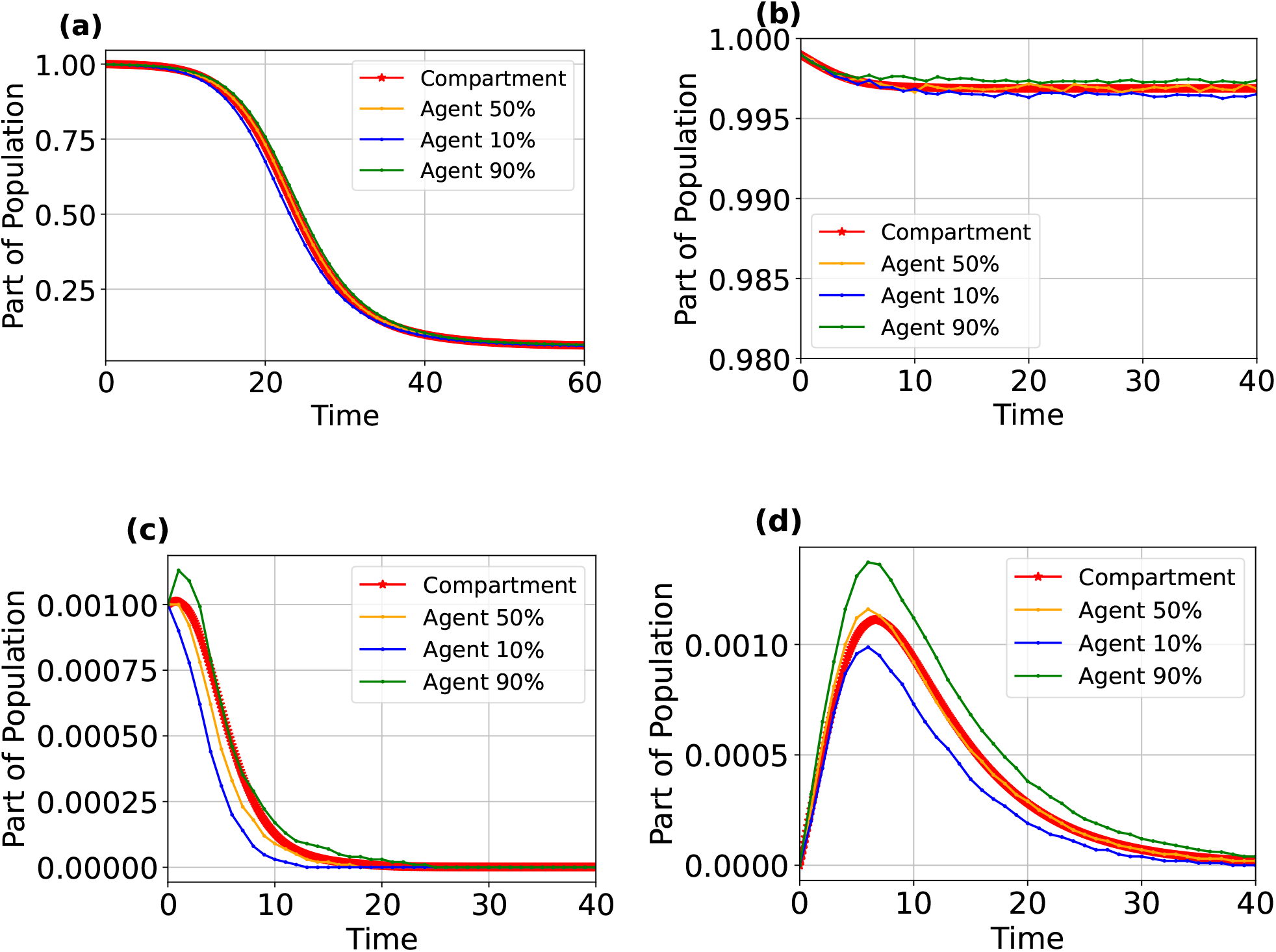
Comparison of the compartment model and agent-based model in the regular-graph case. The sub-figures are susceptible in Newman’s model (a) total susceptible in the SIRCQ model (b), infected and non-quarantined (c), and infected and quarantined (d). The range is between 0 and 1 because we normalize by the population. The parameters of the epidemic are: *β* = 0.002, *γ* = 0.14, *p*_*ct*_ = 0.5, *θ*_*s*_ = 0.07, *p*_*symptoms*_ = 0.25. For (d) symptoms and contact tracing are zero (*p*_*ct*_ = 0, *p*_*symptoms*_ = 0). The degree was chosen to be 281, the same as the mean excess degree in the power-law case.

### Experimental validation of the formula for *R*_0_

The next result validates our theoretical computation of the epidemic threshold. We compare the computed epidemic threshold and the numerical results of the compartmental model. Fig. 4 depicts *r*_∞_ versus the infection parameter (*β*) for power law and regular graphs, respectively, with all other epidemic parameters constant in different scenarios. We observe a good agreement between the theoretical epidemic threshold and the beginning of an exponential growth of the total number of infected people as a function of the infection parameter. In Fig. 4a, since the excess degree is 281, *R*_0_ = 1 for *β* ≈ 0.0014 when quarantine is used and *β* ≈ 0.0005 without quarantine. For the regular graph result, we chose the degree to be the same as the excess degree of the power law graph to have the same thresholds. We can see this result in Fig. 4b.

**Fig. 4:**
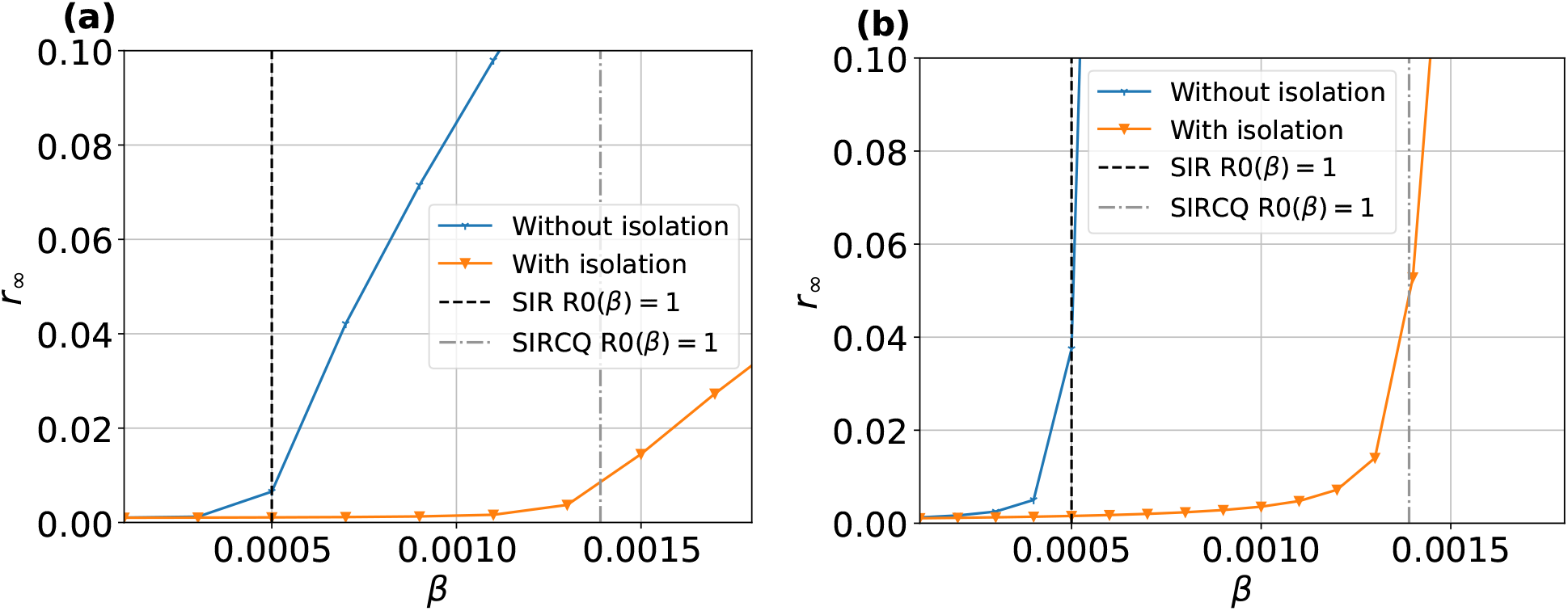
Epidemic threshold expressed with *r*_*∞*_ as a function of *β* in a power law degree distribution(a) and regular-graph degree distribution(b).

### The effects of contact tracing and isolation on the number of infected cases in the population

Our next result studies the impact of contact tracing on the total infected population. We show that accelerating the contact tracing can significantly reduce the total number of infected people. Fig. 5 and Fig. 6 present different scenarios for power law and regular graphs, respectively. Fig. 5a and Fig. 6a show the SIR model with the same degree distribution but without quarantine or contact tracing [43] where there are only three compartments. The range of the infection parameter *β* was selected such that *r*_∞_ covers the interval from 0 to 0.95 in this model. We used the same infection parameters with various contact tracing and quarantine parameters to evaluate the gain of contact tracing and quarantine as compared to this model. Figures 5b and 6b present a different scenario for symptoms-based quarantine only, we can see a significant reduction in the total infection population by up to 30% for the highest probability of infection. The remaining sub-figures consider the effect of adding contact tracing on top of symptoms-based quarantine. We can see a significant reduction in the total infected population, further improving upon the symptoms-based quarantine. In addition, in every scenario, there is a good agreement between the ABM and the compartment model, further validating our model. An important advantage of the compartmental model is that the calculation of these results is 3 orders of magnitude faster compared to the ABM model.

**Fig. 5:**
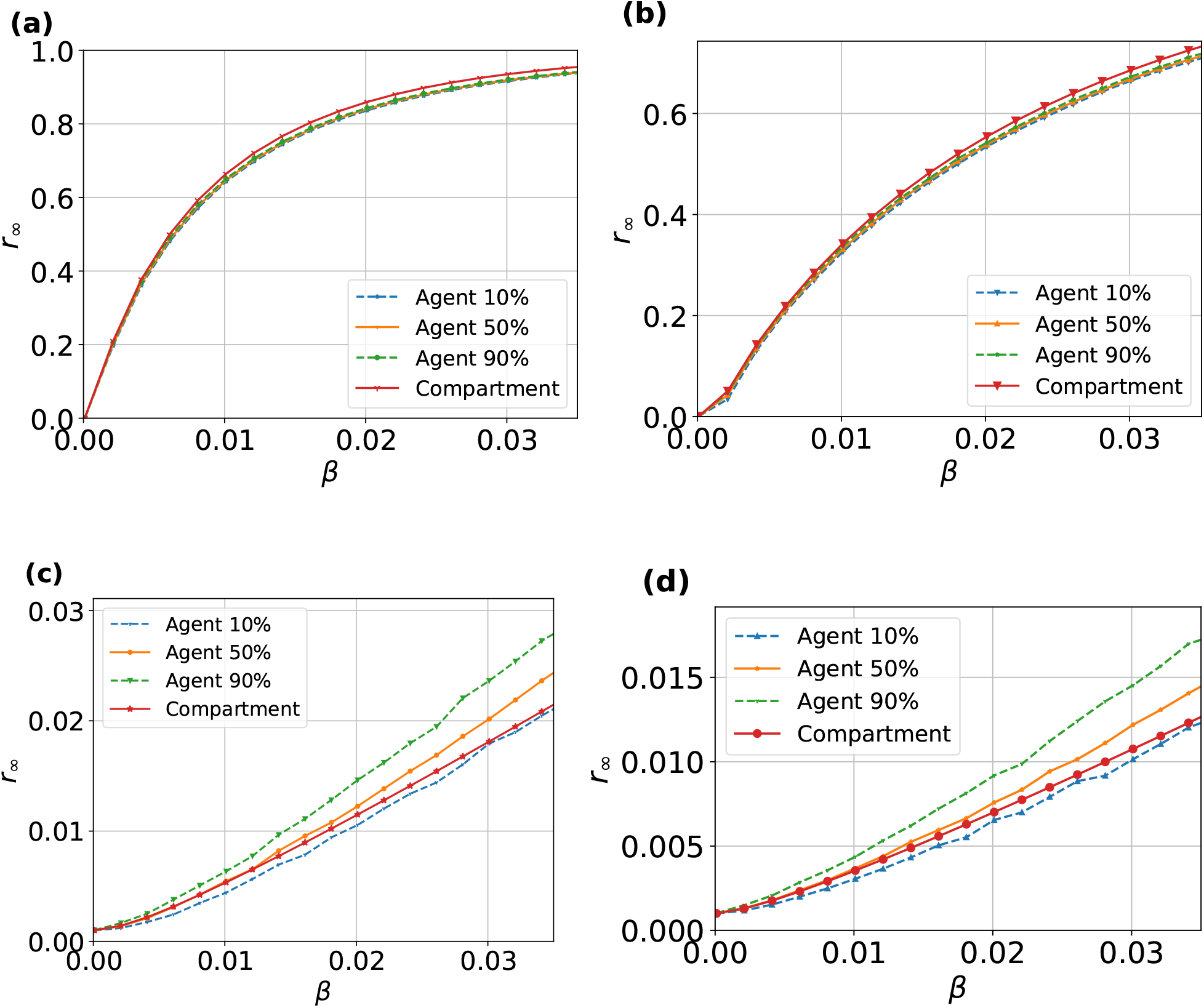
Comparison of *r*_*∞*_ as a function of *β* between agent and compartment in a power law degree distribution. There are 4 sub-figures for different scenarios. Fig. (a) is for No quarantine and no tracing (SIR model with degree distribution), Fig. (b) is for symptom-based quarantine only (*p*_*ct*_ = 0), Fig. (c) is for quarantine and tracing of *p*_*ct*_ = 0.5, Fig. (d) is for quarantine and tracing of *p*_*ct*_ = 0.9.

**Fig. 6:**
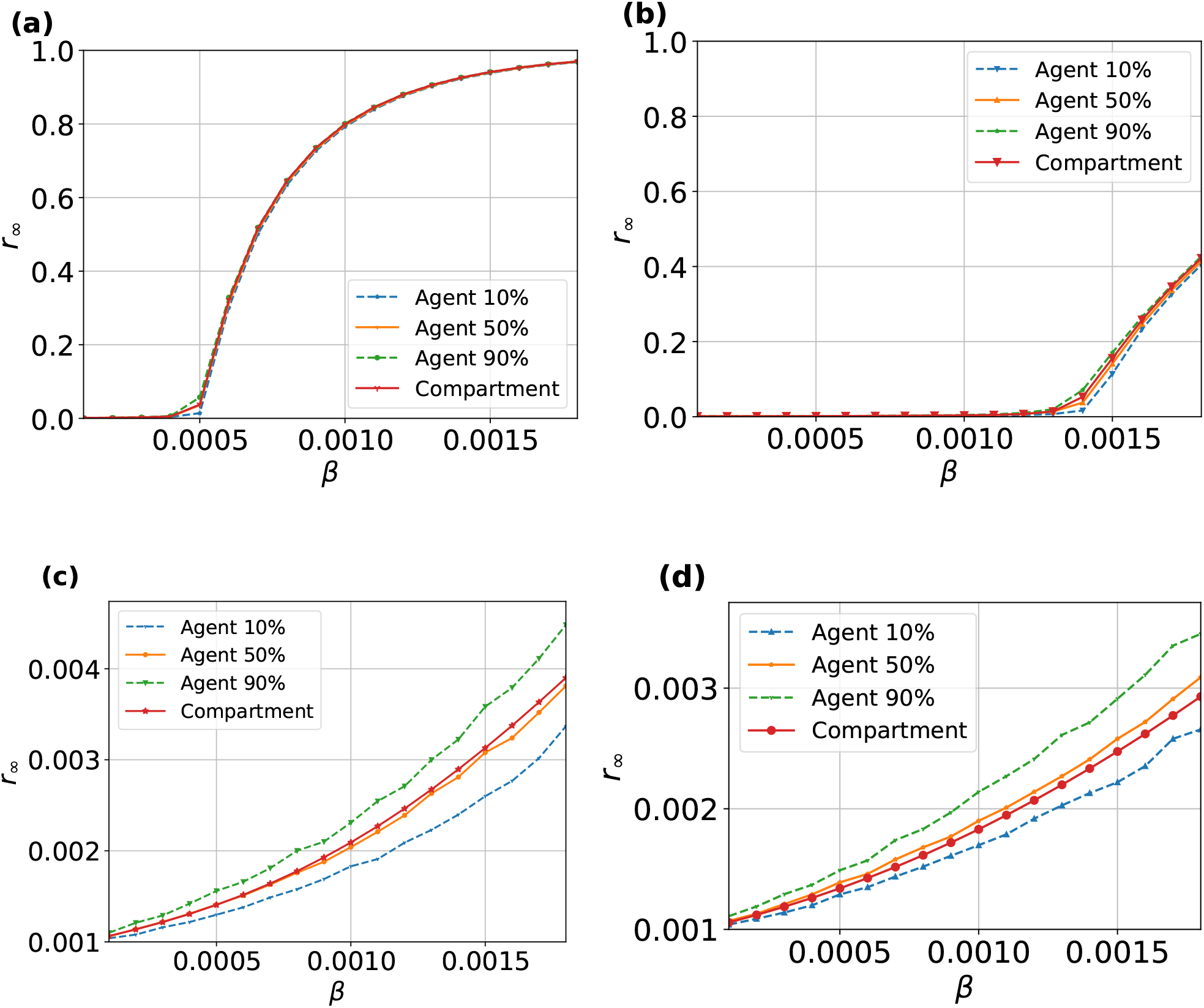
Comparison of *r*_*∞*_ as a function of *β* between agent and compartment in a regular-graph degree distribution. There are 4 traces for different scenarios. Fig. (a) is for No quarantine and no tracing (SIR model with degree distribution), Fig. (b) is for symptom-based quarantine only (*p*_*ct*_ = 0), Fig. (c) is for quarantine and tracing of *p*_*ct*_ = 0.5, Fig. (d) is for quarantine and tracing of *p*_*ct*_ = 0.9. The degree of the graph is 281, as the mean excess degree of the power-law case.

### Comparison between Erdős–Rényi and regular graphs

The differences in the epidemic dynamics between the regular graph and the power law graph are significant. Intuitively, regular graphs represent a case where the variation of the degrees is small. This suggests that the results for regular graphs should be a good approximation to the family of Erdős-Renyi graphs. Our final result is the comparison between Erdős–Rényi and regular graphs. Indeed the results show good agreement over a wide range of parameters. This result is very important because it means that our simplification of the regular graph to a single equation is valid for a wide range of degree distributions which include the Erdős–Rényi graphs. Any degree distribution that can be represented as an Erdős–Rényi graph (or even families where the degree distribution is relatively concentrated) can have a simple approximate solution by numerically solving a single equation with a closed form algebraic solution for the other equations instead of solving numerically 5×*K* equations where *K* is the number of different degrees of nodes. This result is not surprising, as Erdős–Rényi has a rather small variance and is concentrated around the mean. The degree of the regular graph was chosen as the mean of the Erdős–Rényi degree distribution so that we have Erdős–Rényi with *n* as the population size and *p* = *k/n* where k is the degree of the k-regular graph. In Fig. 7 we see the results, we can see that both the agent-based and the compartmental models of Erdős–Rényi are approximately equal to the k-regular graph results.

**Fig. 7:**
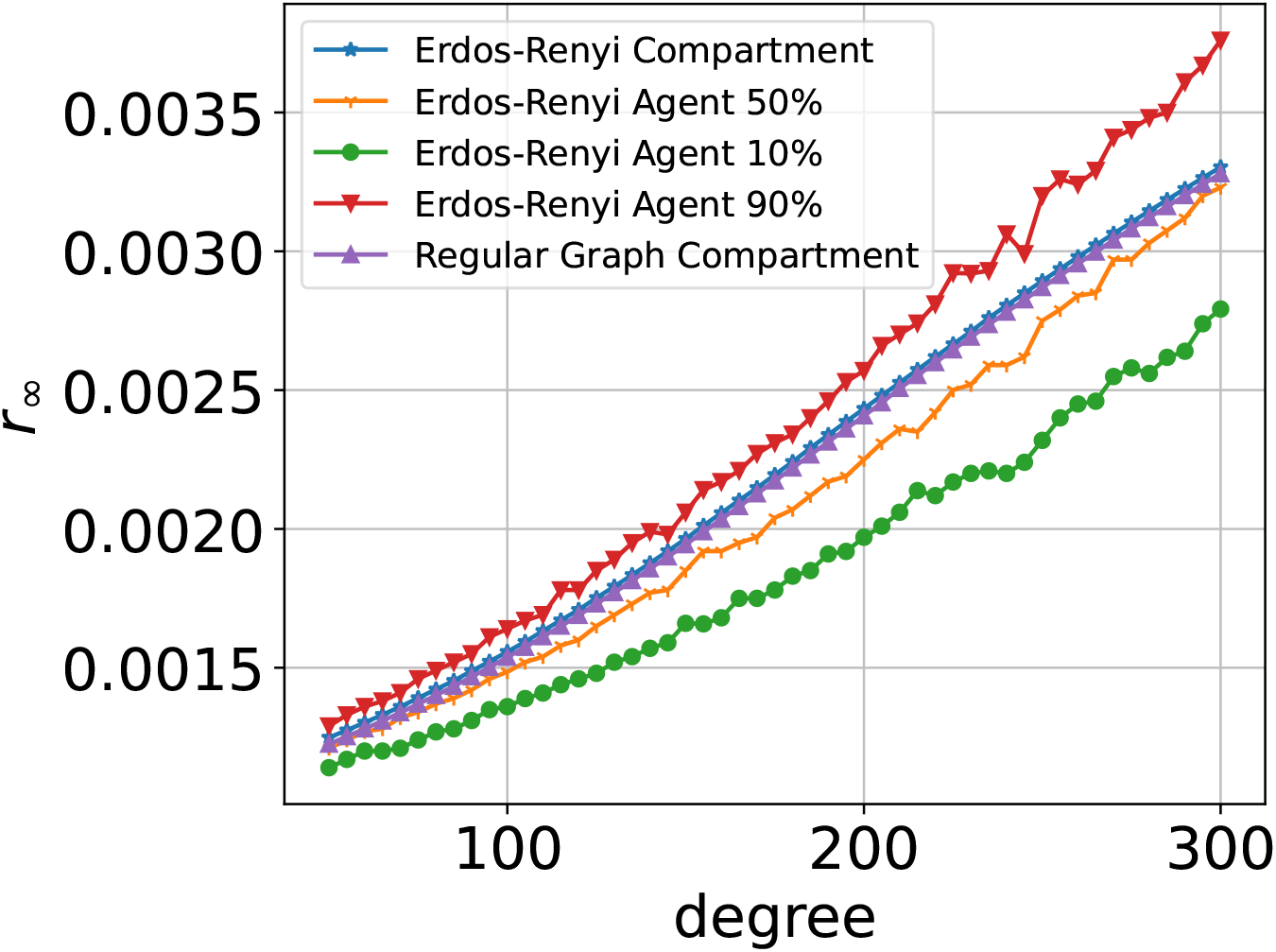
Comparison of *r*_*∞*_ as a function of degree between Erdő s–Rényi and regular graph degree distributions. We have four traces of Erdő s–Rényi, three of which are agent-based simulations and one is compartment-based. We can clearly see that the compartment solution of Erdő s–Rényi agrees with the agent-based solution and that the Erdő s–Rényi compartment solution is almost identical to the regular graph compartment solution. We chose *β* = 0.0015 to make sure *R*_0_ *>* 1 for all degrees in the range.

## Discussion

We introduced a novel compartmental model that includes contact tracing and quarantine under an arbitrary degree distribution. We calculated the epidemic threshold in the general case as well as for regular graphs. Moreover, we developed a way to reduce the complexity of the solution for regular graphs to the numerical solution of a single equation. This model allows a much faster evaluation of the impact of contact tracing and quarantine under various models. Since the solution is 3-4 orders of magnitude faster than the ABM even for populations up to 100,000, the acceleration would be more significant when we try to evaluate the impact of the measures on a state level. Hence, the model provides an important tool for policy making and testing various measures under different situations. It also allows quick sensitivity analysis to the various parameters, something that is much harder with agent-based models, and is important when epidemic parameters are noisy estimates derived from real-time epidemic progression. Furthermore, deriving a simple formula for the epidemic threshold provides an even faster means of estimating the impact of quarantine and contact tracing.

We demonstrated the clear impact of contact tracing on epidemic evolution. Isolation slows the spread of the disease and contact tracing significantly enhances that effect. As a result *r*_∞_ drops.

In addition, we showed that the regular graph case has a good agreement with Erdős–Rényi graphs, which can be used for an entire family of degree distributions. We can further generalize this to a larger family of degree distributions, where the degree variation is small. For example, degree distributions that can be approximated by a two-degree model can be represented by 10 differential equations.

In the future, this model can be a foundation for more complex models, as this model introduces a simple way to incorporate contact tracing into compartmental models.

## Methods

### SIRCQ model development

We now derive the SIRCQ model equations. For each degree, the standard compartments are used for non-quarantined people with degree *k* and denoted by 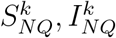. For each degree *k* we add two new compartments for people in quarantine. One for susceptible and quarantined 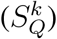 and the other for infected and quarantined 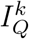. We note that susceptible and quarantined move back to the susceptible and non-quarantined after a certain time, while infected and quarantined move to the recovered compartment (*R*). Each of these transitions has its own rate. The model is depicted in Fig. 1.

To describe the model, we use the following notation: *p*_*k*_ is the probability that a vertex has degree *k, q*_*k*_ that the *excess* degree is *k, x*^*k*^ is the part of the population in the compartment *x*, where *x* is one of the super compartments (i.e, including all degrees) (*s*_*nq*_ - susceptible and not quarantined, *s*_*q*_ - susceptible and quarantined, *i*_*nq*_ - infected and not quarantined, *i*_*q*_ - infected and quarantined, *r* - recovered), normalized by the population size of degree *k*. For example, 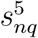 is the proportion of susceptible and not quarantined individuals with degree 5 out of all degree 5 population. *β* is the probability of infection per contact, *γ* is the recovery rate, *v* is the probability that a neighbor vertex is infected and not quarantined (regardless of its degree), 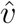 is the probability that a neighbor vertex is infected and quarantined, *p*_*ct*_ is the contact-tracing rate, *p*_*symptoms*_ is the symptoms appearance rate, and *θ*_*s*_ is the rate of exiting quarantine.

The equations for the model are:

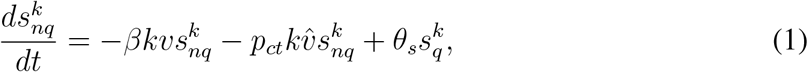

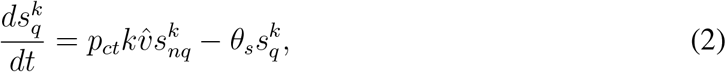

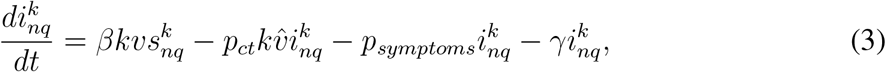

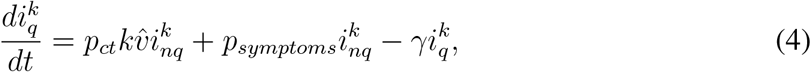

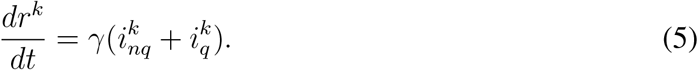

We will now derive these equations similarly to the model of [43]. We begin with the derivation of [12] and modify it to include the new compartments. The equations in [12] are expressed using only 3*K* compartments (*s*_*k*_ - susceptible, *i*_*k*_ - infected, *r*_*k*_ - recovered). Using the notations above the equations in [12] are defined as follows:

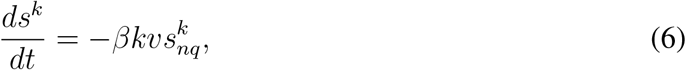

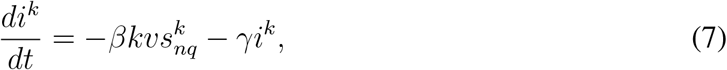

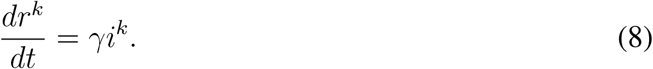

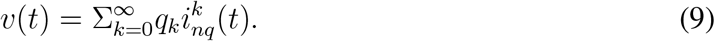

In this model, we have only two transitions: infection (from *s* to *i*) and recovery (from *i* to *r*). In the SIRCQ model, we have a few more transitions: infection (from *s*_*nq*_ to *i*_*nq*_), recovery (from *i*_*nq*_ or *i*_*q*_ to *r*), quarantine from symptoms (from *i*_*nq*_ to *i*_*q*_), contact tracing (from *i*_*nq*_ to *i*_*q*_ or from *s*_*nq*_ to *s*_*q*_), and exit from quarantine (from *s*_*q*_ to *s*_*nq*_). The compartments and the transitions are described in Fig. 1.

In the SIRCQ model the equations (6-9), are modified since the infection can occur only between non-quarantined vertices (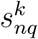and 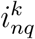), Hence we obtain:

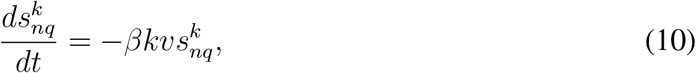

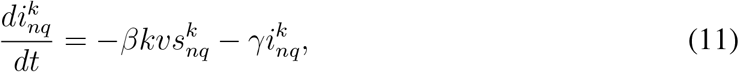

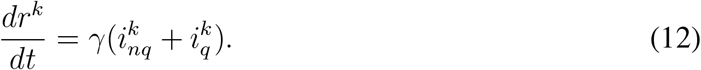

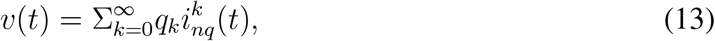

We now include the effect of symptoms. Symptoms occur only in infected people and only transfers people from 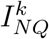 to 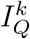. Therefore, equation (11) becomes:

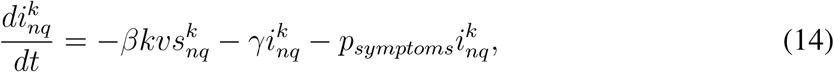

and the new compartment satisfies:

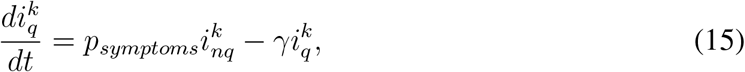

Finally, we address the transition probabilities of the contact tracing. Let us define *p*_*ct*_ as the rate of contact tracing, i.e. the rate of identifying contacts between quarantined people and others. This implies that the contact tracing results in transitions from 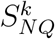 to 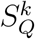 and from 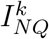 to 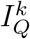. Hence it adds a positive term to 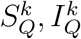 and a negative term to 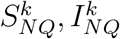. Note that contacts with infected neighbors of any degree impact the contact tracing, and this results in a non-linear term that depends on 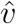 which is defined as the expected excess degree

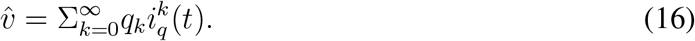

Finally, we arrive at the equations of the model:

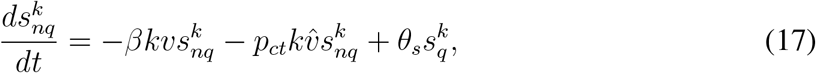

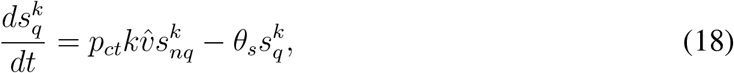

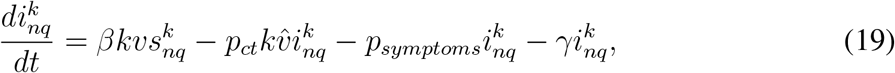

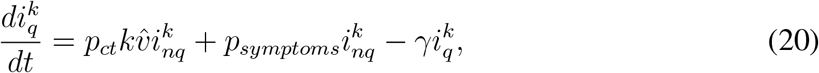

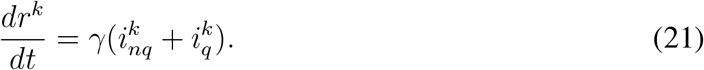

### k-regular graph case

Our system of equations (17-21) does not address the special case of k-regular graph which is in between the classical SIR model and the full arbitrary degree-distribution cases. This specific case is useful, when we know the average number of contacts per person, a parameter which can significantly improve the classical SIR model. In a k-regular graph all vertices have the same degree. Thus, the probability that a neighbor is infected and quarantined or infected and not quarantined is *i*_*q*_ and *i*_*nq*_, respectively. In addition, instead of 5×*K* equations (where *K* is the number of different degrees of nodes), we now only have five equations. Thus, the equations become:

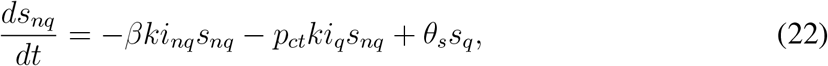

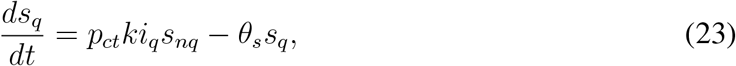

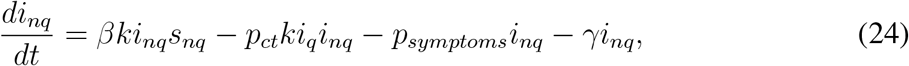

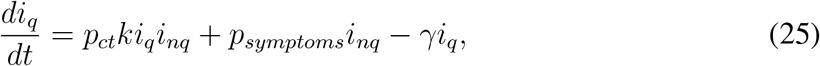

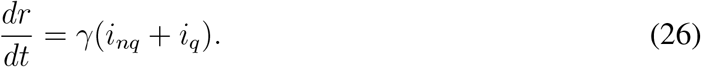

We now show that for *k*-regular graphs, the system (22)-(26) can be reduced into a single equation, as follows:

#### Theorem 1

*Solving the system* (22)*-*(26) *is equivalent to solving the equation*

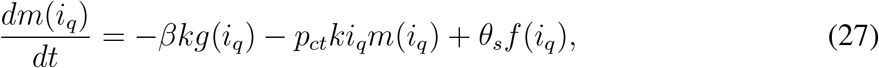

*where:*

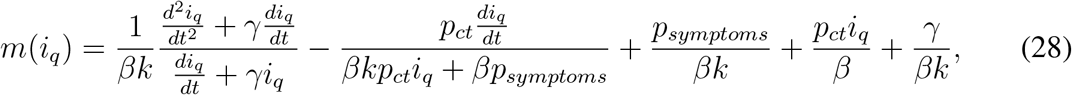

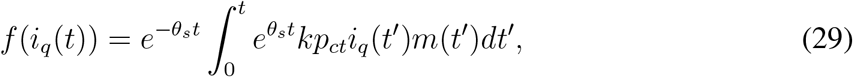

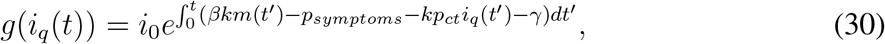

*with the initial conditions: s*_*nq*_(0) = 1−*i*_0_, *i*_*nq*_(0) = *i*_0_, *i*_*q*_(0) = 0, *s*_*q*_(0) = 0, *r*(0) = 0. *All other compartments can be algebraically derived from i*_*q*_(*t*): *s*_*nq*_(*t*) = *m*(*i*_*q*_(*t*)), *s*_*q*_(*t*) = *f* (*i*_*q*_(*t*)), *s*_*nq*_(*t*) = *g*(*i*_*q*_(*t*)) *and r*(*t*) = 1 − *s*_*nq*_(*t*) − *s*_*q*_(*t*) − *i*_*nq*_(*t*) − *i*_*q*_(*t*).

Note that (27) still needs to be solved numerically. However, solving a single differential equation is significantly simpler than solving a system of 5 non-linear differential equations.

*Proof*. To prove the theorem we can first solve the ODE (24) for *i*_*nq*_ with the initial condition *i*_*nq*_(0) = *i*_0_ obtaining:

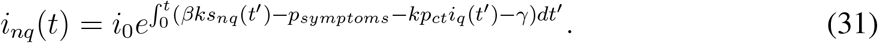

We now substitute equation (31) with *i*_*nq*_ in equation (25). We do this in order to find a solution for *s*_*nq*_ so that *s*_*nq*_ = *f* (*i*_*q*_):

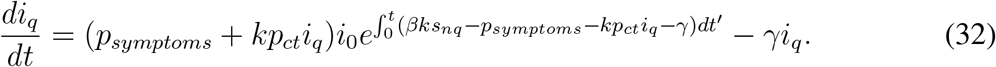

We isolate the exponent and use the natural logarithm on both sides:

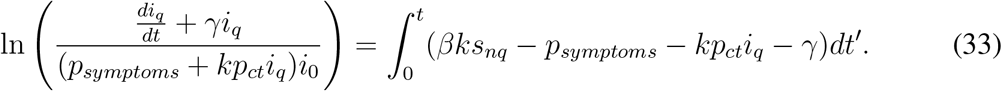

We now derive both sides and isolate *s*_*nq*_ to get:

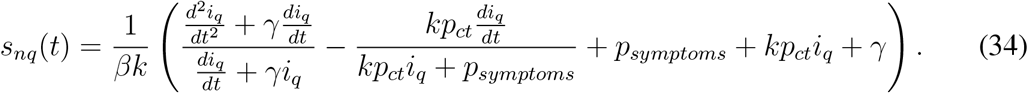

Afterward, we solve the ODE in equation (23) for *s*_*q*_ with the initial condition that *s*_*q*_(0) = 0:

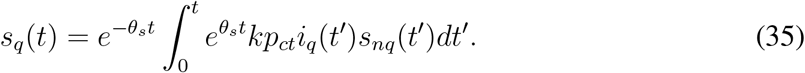

Since we can express *s*_*nq*_, *s*_*q*_, *i*_*nq*_ as functions of *i*_*q*_, we can substitute equations (35, 34, 31) to equation (22) to get an ODE of *i*_*q*_. In order to simplify the equation, we will use the following notations:

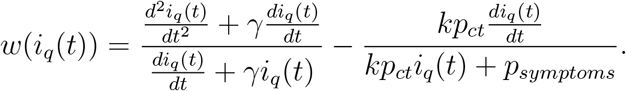

The ODE equation of *i*_*q*_ is:

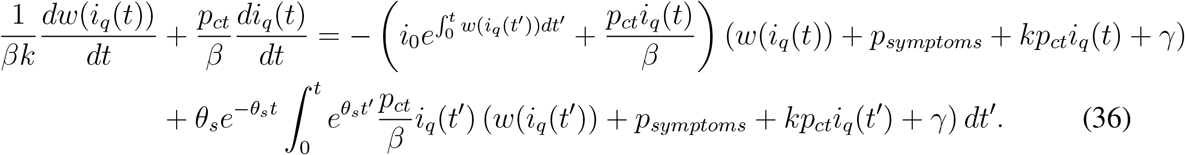

Once we solve the ODE of *i*_*q*_ numerically, we can calculate the rest of the compartments using equations (35, 34, 31).

### Calculation of the epidemic threshold with an arbitrary degree distribution

#### Theorem 2

*The epidemic threshold is* 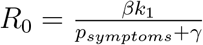.

*Proof*. We use the next generation matrix method from [42] to calculate *R*_0_.The infectious relevant compartments are 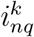 and 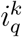. Their subsystem of equations is:

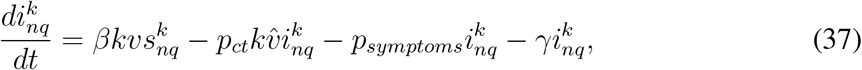

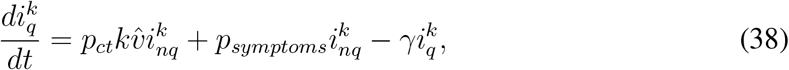

for 1 ≤ *k* ≤ *n*, where 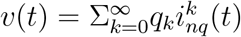 and 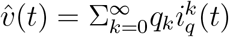.

The DFE (disease-free equilibrium) of our system of equations (17–21) is 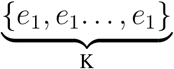, where *K* is the number of the degrees and *e*_1_ = (1, 0, 0, 0, 0). Meaning 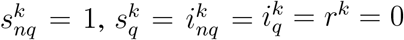 for all *k*.

We will first derive equations 37, 38 by 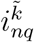 for each 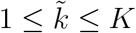:

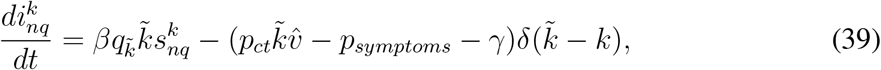

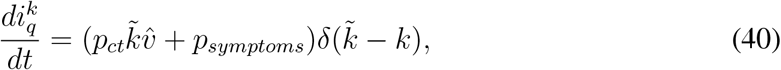

where 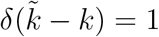 for 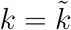 and 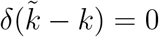 otherwise. Equations 39, 40 represent 2*K* equations for (2 for each 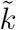).

We will do the same derivation but by 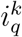 this time:

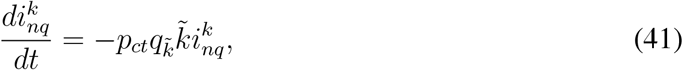

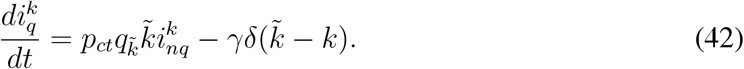

Afterward, we will substitute the DFE and represent equations 39, 40, 41, 42 in a matrix form for all *K* degrees.

The *K* × 2*K* Jacobian matrix is represented as:

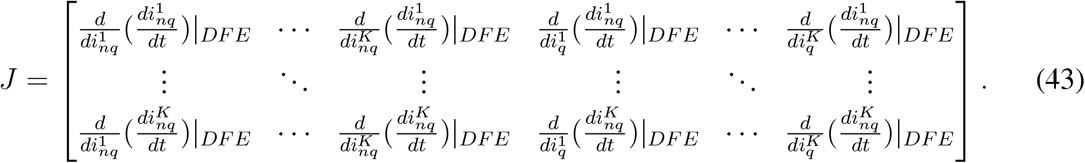

If we compute *J* we get:

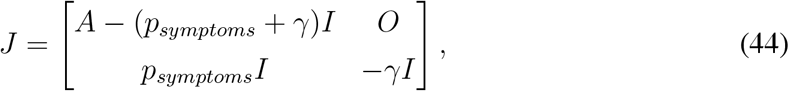

where *I* and *O* are the identity and zero matrix of order *K*, respectively, and the matrix A is:

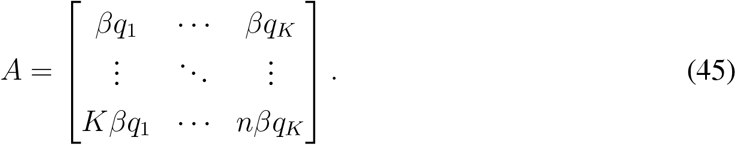

The Jacobian matrix is decomposed into a transmission matrix *F* and transition matrix *V*:

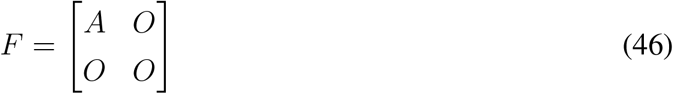

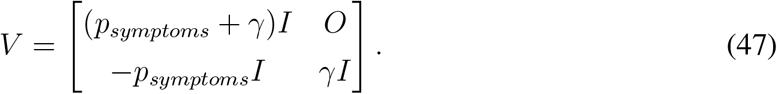

The inverse of the transition matrix is:

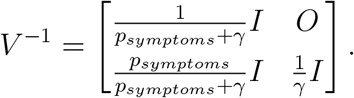

Next, we find the spectral radius of the next generation matrix *FV* ^−1^:

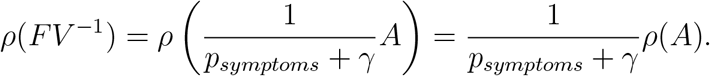

Using Lemma 3 we obtain:

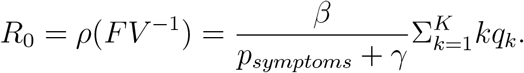

Finally, we get:

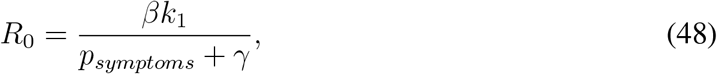

where *k*_1_ is the mean excess degree.

#### Lemma 3

*The spectral radius of A is* 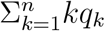

*Proof*. Let the vectors **a, b** be defined by:

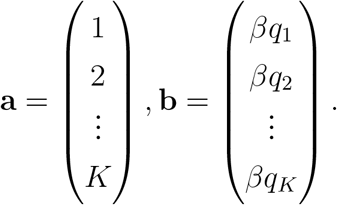

We can clearly see that *A* = **ab**^*T*^. The matrix *A* has rank 1 since all the rows of *A*’s are linearly dependent. Hence there is a single positive eigenvalue which is **a** since

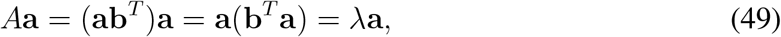

Where 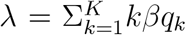 is the positive eigenvalue. Because *A* has only a single non-zero eigen-value, *λ* is also the maximal eigenvalue and thus the spectral radius.

#### Calculation of the epidemic threshold for k-regular graph

In the *k*-regular case the epidemic threshold can be derived in a simpler more intuitive way by considering equation (24). An epidemic is spreading only when the factor *βks*_*nq*_ − *p*_*symptoms*_ − *kp*_*ct*_*i*_*q*_ − *γ* is positive. In early phase of the epidemic *s*_*nq*_ ≈ 1 and *i*_*q*_ ≈ 0, therefore, equation (24) becomes:

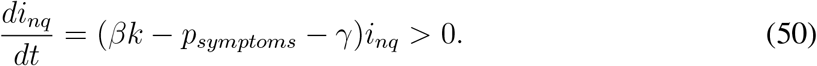

Thus an outbreak occurs when

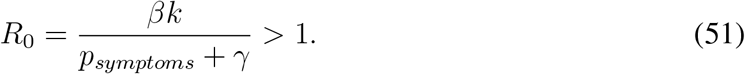

We can also use the next-generation matrix method to get the same result, where the regular graph case is easier than the general degree distribution.

### Numerical solution of the equations

In order to solve the differential equations numerically, we used the explicit Runge-Kutta method of order 5(4) [44] as our integration method. We used a single vector of all compartments for the calculation, i.e.:

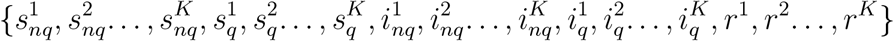

with the initial conditions:

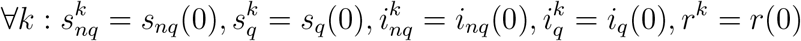

The initial conditions stay the same for each degree to get the correct initial condition for the corresponding compartment. For example, for susceptible and not quarantined:

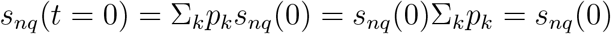

### Agent-based model

In order to verify the model, we developed simulations of an agent-based model. The simulation results are then compared to the predicted results of the SIRCQ model.

*Population network*: In our simulations, we use a population of *n* = 10^5^ and a configuration model to attach the proper degree distribution to the vertices upon each realization. We used random graphs with a power law degree distribution as well as regular graphs with varying degrees. The reasoning for using a power-law degree distribution is that many real networks, such as contact networks, have been shown to exhibit scale-free properties as mentioned in [45]. Regular graphs are special cases with a single degree. This model generalizes the classical SIR, where each node has only a relatively small number of possible contacts.

The probability *P* (*k*) that a node is connected to k other nodes follows a Power-law:

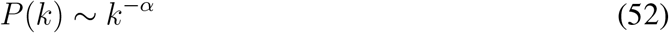

where *α* is usually ranging between 2 ≤ *α* ≤ 3. We used the following parameters:

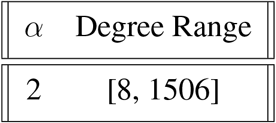

We chose a small *α* and the degree range to have a slower and more realistic fade of degrees. *Epidemic parameters*: The parameters we chose for the epidemic unless noted otherwise are

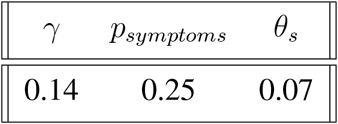

The reason we chose these parameters is to have a mean time of recovery of 7 days, mean time of symptoms appearance of 4 days and quarantine time of 14 days. Because the rates of *γ, p*_*symptoms*_, *θ*_*s*_ act like a geometric distribution, the rates need to be *1/(mean time)*.

## Data Availability

All data produced in the present study are available upon reasonable request to the authors

